# Effectiveness of the adjuvanted Sanofi/GSK (VidPrevtyn Beta) and Pfizer-BioNTech (Comirnaty Original/Omicron BA.4-5) bivalent vaccines against hospitalisation amongst adults aged 75 years and older in England, estimated using a test-negative case control study design

**DOI:** 10.1101/2023.09.28.23296290

**Authors:** Freja Cordelia Møller Kirsebom, Nick Andrews, Julia Stowe, Gavin Dabrera, Mary Ramsay, Jamie Lopez Bernal

## Abstract

**Background:** In England, the Joint Committee for Vaccination and Immunisation recommended a spring 2023 booster programme for all adults aged 75 years and older and the immunosuppressed. The vaccines advised were the Sanofi/GSK AS03-adjuvanted monovalent beta variant (VidPrevtyn Beta) booster vaccine and the Pfizer-BioNTech mRNA (Comirnaty Original/Omicron BA.4-5) bivalent vaccine. This is the first time an adjuvanted COVID-19 vaccine has been administered as part of a UK COVID-19 vaccination programme. In clinical trials, the antibody levels generated by the Sanofi/GSK vaccine were comparable to levels generated by COVID-19 mRNA vaccines but to date there are no real-world data on the effectiveness or duration of protection of this vaccine.

**Methods:** We used a test-negative case-control study design to estimate the incremental vaccine effectiveness of the Sanofi/GSK and Pfizer bivalent BA.4-5 boosters against hospitalisation amongst those aged 75 years and older in England. The study period for tests contributing to all analyses was from 3rd April 2022 to 27th August 2023. Vaccine effectiveness was estimated relative to those who had received at least two doses prior to their spring booster, with their last dose being an autumn 2022 booster given at least 3 months prior.

**Findings:** Overall, there were 14,174 eligible tests from hospitalised individuals aged 75 years and older, with 3,005 being cases and 11,169 being controls. Effectiveness against hospitalisation was highest in the period 9 to 13 days post vaccination for both manufacturers at about 50%; 43.6% (95% C.I.; 20.1 to 60.2%) and 56.4% (95% C.I; 25.8 to 74.4%) for Sanofi/GSK and Pfizer BA.4-5, respectively. There was some evidence of waning with a reduction to about 30% for both manufacturers 5-9 weeks post vaccination.

**Interpretation:** Together, these results provide reassuring evidence that both the adjuvanted Sanofi/GSK and Pfizer BA.4-5 booster vaccines provided a good boost to protection against hospitalisation amongst adults aged 75 years and older. The finding that the adjuvanted vaccine targeting the now distant Beta strain had similar effectiveness to the mRNA vaccine targeting more closely matched Omicron sub-lineages BA.4-5 during a period of Omicron circulation may reflect improved protection due to the adjuvant in the Sanofi/GSK product.

**Funding:** No external funding.

## Introduction

COVID-19 vaccines provide good protection against severe disease, (1-5) however, due to waning of the vaccines, many countries have opted for frequent booster programmes to maintain a high level of protection in the most vulnerable populations. COVID-19 continues to evolve and over the past 18 months sub-lineages of Omicron have emerged including BA.1, BA.2 (6), BA.4, BA.5 (7), BQ.1, CH.1.1, numerous sub-lineages related to XBB (8, 9) and most recently BA.2.86 (10, 11). Manufacturers have developed modified vaccines which target the spike protein on different COVID-19 variants.

COVID-19 booster vaccinations were offered in the UK in autumn 2021 (all adults), spring 2022 (over 75 year olds and immunosuppressed individuals), autumn 2022 and spring 2023 (12). The autumn 2022 booster programme commenced the 5^th^ September 2022 (13) and bivalent BA.1 boosters with either Pfizer BioNTech (Original/Omicron BA.1 Comirnaty®) or Moderna (Spikevax® bivalent Original/Omicron BA.1 vaccine) were offered to all adults aged 50 years and over and vulnerable individuals (14). The spring 2023 booster programme targeted all adults aged 75 years and older, and the immunosuppressed (15). The vaccines advised included the Sanofi/GSK AS03-adjuvanted monovalent beta variant (VidPrevtyn Beta) booster vaccine, the Pfizer-BioNTech mRNA (Comirnaty Original/Omicron BA.4-5) bivalent vaccine and the Moderna mRNA (Spikevax bivalent Original/Omicron BA.4-5) bivalent vaccine. The programme commenced from 3^rd^ April 2023, with care home residents being prioritised for vaccination. The Sanofi/GSK booster was rolled out first, followed by Pfizer BA.4-5 and Moderna BA.4-5.

This is the first time the Sanofi/GSK vaccine has been administered as part of a UK COVID-19 vaccination programme, and the first time an adjuvated COVID-19 vaccine has been used in the UK. It is a protein sub-unit vaccine containing the beta spike protein in combination with the AS03 adjuvant (15). This vaccine was approved by the Medicines and Healthcare products Regulatory Agency (MHRA) in the UK for use in adults (aged over 18 years) who have already received an mRNA or adenoviral vector COVID-19 vaccine. Adjuvanted vaccines against influenza have been shown to have higher effectiveness and longer duration of protection (16, 17). In clinical trials, the antibody levels generated against different Omicron sub-variants by the Sanofi/GSK vaccine were comparable to levels generated by COVID-19 mRNA vaccines (18).

Here, we used a test-negative case control (TNCC) study design to estimate vaccine effectiveness against hospitalisation amongst those aged 75 years and older of the Sanofi/GSK and Pfizer BA.4-5 boosters administered as part of the spring 2023 booster programme in England. During this period, XBB-related sub-lineages dominated in England (9).

## Methods

### Study design

To estimate VE against hospitalisation, a TNCC study design was used where all positive PCR tests from individuals aged 75 years and older hospitalised with a respiratory code in the primary diagnosis field in England are cases while all negative PCR tests from such individuals are controls, as previously described (1-5).

### Data Sources

#### COVID-19 Testing Data

Cases and controls were identified from national COVID-19 PCR testing data performed in hospital settings (Pillar 1). The study period included all positive and negative Pillar 1 PCR tests from 3^rd^ April 2022 to 27^th^ August 2023.

Negative tests taken within 7 days of a previous negative test were dropped as these likely represent the same episode. Negative tests taken within 21 days of a subsequent positive test were also excluded as chances are high that these are false negatives. Tests within 90 days of a previous positive test were also excluded as these likely represent the same episode. The date of an individual’s most recent prior positive test was identified from all historic testing data (Pillar 1 or Pillar 2). Individuals contributed a maximum of one negative control test. Further details on the reasons for these exclusions are described previously (19).

### National Immunisation Management System (NIMS)

The National Immunisation Management System (NIMS) is a national vaccine register containing demographic information on the whole population of England registered with a GP, used to record all COVID-19 vaccinations (20).

Testing data were linked to NIMS using combinations of the unique individual NHS number, date of birth, surname, first name, and postcode using deterministic linkage. NIMS was accessed for dates of vaccination and manufacturer, sex, date of birth, ethnicity, care home residency and residential address. Care home status is provided by the NHS. We identified those being resident in a care home as of March 2023. Addresses were used to determine index of multiple deprivation quintile. Data on risk group status (those identified as at risk previously in the pandemic and those identified recently as requiring an autumn booster by NHS CaaS (Cohorting as a Service) (21)), clinically extremely vulnerable status, severely immunosuppressed and health/social care worker status were also extracted from the NIMS. Booster doses given as part of the autumn 2022 and spring 2023 booster programmes were classified based on SNOMED coding and timing (autumn booster doses were those coded as bivalent BA.1 doses (Moderna or Pfizer) administered from 5^th^ September 2022, spring booster doses were those coded as bivalent BA.4-5 (Moderna or Pfizer) or Sanofi/GSK administered from 3^rd^ April 2023).

Only individuals who had received at least two doses (a primary vaccination course) prior to the 3^rd^ April 2023 and whose last dose prior to the 3^rd^ April was given as part of the autumn booster programme were included. Those with less than a 12-week interval between the autumn booster dose and the spring booster dose and those who received more than one dose coded as a spring booster dose were excluded. Additionally, those who received a booster dose after the 3^rd^ April 2023 which wasn’t a bivalent BA.4-5 (Moderna or Pfizer) or Sanofi/GSK dose were excluded.

### Hospital Admission Data

Secondary Uses Service (SUS) is the national electronic database of hospital admissions that provides timely updates of ICD-10 codes for completed hospital stays for all NHS hospitals in England (22). Hospital inpatient admissions for a range of acute respiratory illnesses (ARI) were identified from the SUS and linked to the testing data using NHS number and date of birth. Length of stay was calculated as date of discharge minus the date of admission. Admissions were restricted to those with an ICD-10 coded acute respiratory illness (ARI) discharge diagnosis in the primary diagnosis field (Supplementary Table 1), who were admitted for at least 2 days, where the date of test was 1 day before and up to 2 days after the admission.

**Table 1.** Vaccine effectiveness (VE) against hospitalisation amongst those aged 75 years and older in England, stratified by spring booster manufacturer.

| Spring booster* | Interval | Controls | Cases | Odds Ratio | VE (95% C.I.) |
| --- | --- | --- | --- | --- | --- |
| None | - | 6,334 | 2,150 | Baseline | Baseline |
| Sanofi | 0-2 days | 82 | 8 | 0.30 (0.15-0.63) | 69.6 (36.6 to 85.5) |
|  | 3-8 days | 215 | 63 | 0.84 (0.63-1.14) | 15.5 (-13.8 to 37.3) |
|  | 9-13 days | 212 | 41 | 0.56 (0.4-0.8) | 43.6 (20.1 to 60.2) |
|  | 2-4 weeks | 808 | 156 | 0.67 (0.55-0.81) | 33.2 (18.9 to 45.0) |
|  | 5-9 weeks | 1015 | 131 | 0.67 (0.54-0.83) | 33.3 (17.0 to 46.4) |
|  | 10+ weeks | 855 | 203 | 0.82 (0.65-1.03) | 17.7 (-3.5 to 34.6) |
| Pfizer BA.4-5 | 0-2 days | 61 | 9 | 0.49 (0.24-1.0) | 51.3 (0.3 to 76.2) |
|  | 3-8 days | 132 | 27 | 0.76 (0.49-1.16) | 24.4 (-16.0 to 50.8) |
|  | 9-13 days | 136 | 16 | 0.44 (0.26-0.74) | 56.4 (25.8 to 74.4) |
|  | 2-4 weeks | 483 | 52 | 0.47 (0.35-0.64) | 52.9 (36.2 to 65.2) |
|  | 5-9 weeks | 567 | 86 | 0.71 (0.54-0.92) | 29.5 (7.9 to 46.0) |
|  | 10+ weeks | 269 | 63 | 0.62 (0.44-0.87) | 37.8 (13.1 to 55.5) |

\*All individuals had received a bivalent BA.1 booster vaccine as part of the autumn 2022 booster programme, and their last dose was at least 3 months prior to their test.

### Covariates and adjustment

Potential confounding variables were week of test date (categorical), gender, age (five-year age bands), risk group status, care home status, health and social care worker status, region, IMD quintile, ethnicity, influenza vaccination status this season and the likely variant of an individual’s most recent prior infection.

### Statistical methods

Multivariable logistic regression was used with the test result as the outcome, vaccination status as the primary variable of interest and with confounder adjustment as described above. VE was calculated as 1-odds ratio and given as a percentage. Estimates were not shown where there were less than 30 controls.

In recent studies of vaccine effectiveness by our group and others, we have estimated effectiveness as the extra protection of a booster dose in addition to the protection already provided by previous vaccinations (5, 8, 23, 24). Since most of the adult population in England has now received multiple COVID-19 vaccine doses as part of primary and booster vaccination campaigns, very few individuals remain unvaccinated. We consider it most relevant therefore to estimate the additional benefit of a booster vaccine in addition to the protection an individual has from past doses which have waned in effectiveness. Thus, we here estimated the incremental vaccine effectiveness of the spring boosters amongst those who had received a bivalent BA.1 booster dose as part of the autumn 2022 programme, where the previous dose was given at least 3 month prior. The effectiveness estimated here is therefore the additional protection of the spring booster in addition to the protection conferred by at least two doses where the last dose received was a bivalent BA.1 booster.

Incremental VE was estimated at the following intervals since booster vaccination; 0 to 2 days, 3 to 8 days, 9 to 13 days, 2 to 4 weeks, 5 to 9 weeks and 10 or more weeks. We split the data 0 to 2 and 3 to 8 days to distinguish any very early protective effect from the deferral of vaccination in individuals who were already aware of their COVID-19 positive status from home lateral flow testing or a healthy vaccinee effect in which those with early symptoms of COVID-19 were more unwell than controls and were less likely to be vaccinated from the period 3 to 8 days where we do not expect to see a true protective effect from vaccination. Analyses were stratified by manufacturer. Additionally, sensitivity analyses were conducted restricting to those who were not resident in a care home as the initial roll-out of the vaccines was primarily with the Sanofi/GSK boosters with residents of care homes being prioritised for vaccination. The effect not adjusting for prior infection was also assessed.

## Results

Overall, there were 14,174 eligible tests from hospitalised individuals aged 75 years and older, with 3,005 being cases and 11,169 being controls. Full descriptive characteristics are available in Supplementary Table 2. The distribution of cases and controls over time is shown in Supplementary Figure 1 and the distribution of time since spring booster vaccination by manufacturer is shown in Supplementary Figure 2.

**Table 2.** Vaccine effectiveness (VE) against hospitalisation amongst those aged 75 years and older not resident in a care home, stratified by spring booster manufacturer.

| Spring booster* | Interval | Controls | Cases | Odds Ratio | VE (95% C.I.) |
| --- | --- | --- | --- | --- | --- |
| None | - | 5,902 | 2,036 | Baseline | Baseline |
| Sanofi | 0-2 days | 51 | 5 | 0.27 (0.11-0.68) | 73.1 (31.8 to 89.3) |
|  | 3-8 days | 158 | 54 | 0.90 (0.65-1.25) | 9.9 (-24.7 to 34.9) |
|  | 9-13 days | 153 | 29 | 0.49 (0.32-0.74) | 51.1 (26.1 to 67.6) |
|  | 2-4 weeks | 630 | 132 | 0.68 (0.55-0.84) | 31.7 (15.6 to 44.7) |
|  | 5-9 weeks | 789 | 111 | 0.68 (0.53-0.86) | 32.4 (14.4 to 46.6) |
|  | 10+ weeks | 640 | 181 | 0.85 (0.67-1.08) | 15.0 (-8.2 to 33.1) |
| Pfizer BA.4-5 | 0-2 days | 55 | 7 | 0.40 (0.18-0.9) | 59.5 (9.7 to 81.9) |
|  | 3-8 days | 114 | 25 | 0.78 (0.5-1.22) | 21.9 (-22.4 to 50.2) |
|  | 9-13 days | 121 | 15 | 0.45 (0.26-0.78) | 55.2 (22.2 to 74.2) |
|  | 2-4 weeks | 447 | 51 | 0.49 (0.36-0.67) | 50.9 (33.1 to 63.9) |
|  | 5-9 weeks | 525 | 83 | 0.70 (0.53-0.92) | 29.8 (7.7 to 46.6) |
|  | 10+ weeks | 241 | 60 | 0.61 (0.43-0.86) | 38.8 (13.7 to 56.6) |

\*All individuals had received a bivalent BA.1 booster vaccine as part of the autumn 2022 booster programme, and their last dose was at least 3 months prior to their test.

**Figure 1.**
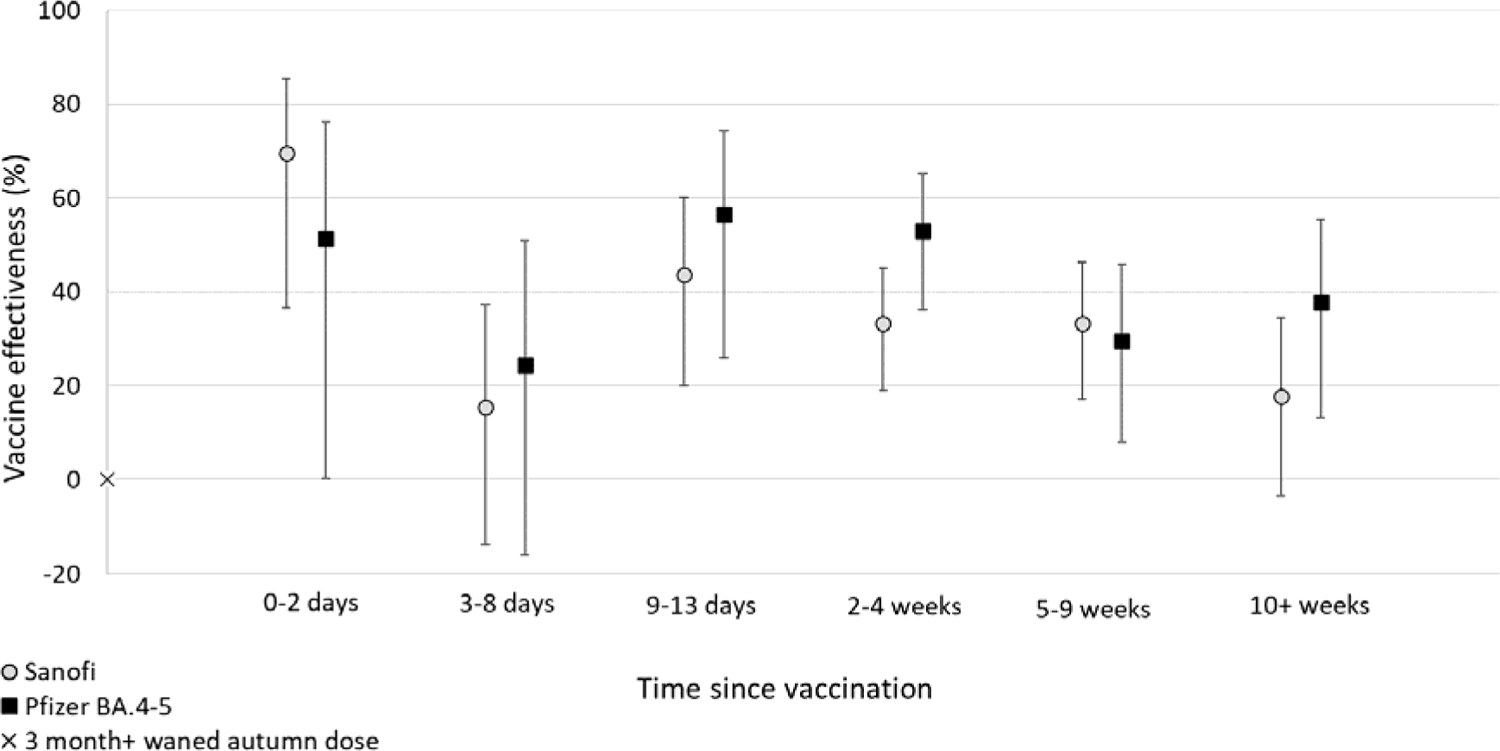
Vaccine effectiveness (VE) against hospitalisation amongst those aged 75 years and older in England, stratified by spring booster manufacturer.

Odds ratios and corresponding vaccine effectiveness estimates are given in Table 1 and Figure 1. The odds of vaccination by Sanofi/GSK and by Pfizer BA.4-5 just prior to testing (0 to 2 days) was significantly below one, yielding apparent positive vaccine effectiveness. The odds increased to closer to one and were non-significant in the 3 to 8 day period indicating no vaccine association, as may be expected in a period before vaccine induced immunity is expected. From 9 days after vaccination, when true vaccine protective effects may be expected, VE against hospitalisation was highest in the earliest period 9 to 13 days post vaccination for both manufacturers at about 50%. From 2 to 4 weeks VE the point estimate was slightly lower for Sanofi/GSK at 33.2% (95% C.I.; 18.9 to 45.0%) compared to Pfizer BA.4-5 booster at VE 52.9% (95% C.I.; 36.2 to 65.2%) though confidence intervals overlapped. After 5 to 9 weeks, VE was somewhat lower than at 9 to 13 days post vaccination for both manufacturers at 33.3% (95% C.I.; 17.0 to 46.4%) for the Sanofi/GSK booster and 29.5% (95% C.I.; 7.9 to 46.0%) for the Pfizer BA.4-5 booster. From 10 weeks post vaccination there was some non-significant evidence of a further decline for Sanofi/GSK but not Pfizer BA.4-5, although confidence intervals were wide and overlapping between the manufacturers and within this period the interval since vaccination will have been longer for Sanofi/GSK as more of this vaccine was used at the start of the campaign.

When the analysis was restricted to only include individuals who are not currently living in a care home, VE estimates against hospitalisation for the Sanofi/GSK booster and Pfizer BA.4-5 booster were similar to those obtained using all cases and controls (Table 2). In sensitivity analyses where the adjustment for past positivity was removed, point estimates differed from the primary analysis by less than 2% in the time intervals from 9 days post vaccination.

## Discussion

In this test-negative case-control study, we estimated the protection conferred by the Sanofi/GSK (VidPrevtyn Beta) and Pfizer BA.4-5 booster vaccines against hospitalisation amongst those aged 75 years and older in England – the main target group of the COVID-19 spring booster campaign due to the increased vulnerability of older adults to severe COVID-19. During the study period, XBB sub-lineages dominated in England. We found that the incremental vaccine effectiveness of a booster dose with either vaccine peaked at around 50% protection against hospitalisation in addition to any remaining protection from prior vaccination with the bivalent BA.1 booster given as part of the autumn 2022 programme.

To our knowledge these are the first real-world effectiveness data for an adjuvanted COVID-19 vaccine. Immunogenicity studies have found that the Sanofi/GSK vaccine elicited as high or higher neutralizing-antibody titres as the original Pfizer mRNA vaccine did against the original strain and against the Beta, Delta, and Omicron BA.1 variants when these vaccines were given as booster doses (25). Unpublished data from the UKHSA CONSENSUS study found antibody responses to be broadly similar between the two vaccines. The efficacy of an adjuvanted bivalent vaccine (ancestral/Beta) by Sanofi/GSK against symptomatic COVID-19 was 75% in SARS-CoV-2 non-naive participants when given as a second dose in a clinical trial (26).

Overall, we found both vaccines provided good protection against hospitalisation (around 30-50% in addition to the protection from a waned autumn booster) and we did not find significant differences in the effectiveness of the boosters by manufacturer. The most prevalent strains in the UK during this analysis period were XBB sub-lineages which are more closely related to the BA.4/5 lineages which this mRNA bivalent vaccine is based on. The similar level of protection with the more distant Sanofi/GSK vaccine based on the Beta strain may be attributable to the adjuvant in the Sanofi product, the different vaccine platform or the use of bivalent mRNA vaccine, where the immune system may be better directed against the original strain rather than the second target strain. Further comparisons of real-world protection from different vaccine platforms and formulations in the presence of matched and unmatched strains are warranted.

Other studies have assessed the real-world effectiveness of the Pfizer BA.4-5 bivalent booster vaccine and found broadly similar results to us; one study from the United States found the incremental effectiveness against hospitalisation of a BA.4-5 bivalent booster was 42% when the last dose was monovalent and given 5 to 7 months prior (23), while another found the relative effectiveness against hospitalisation was 52% when the last dose was monovalent (27). Meanwhile a recent Italian study found the relative effectiveness of bivalent BA.4-5 boosters was 50.6% against severe disease when the last dose was given at least 120 days prior (28). Although the Moderna bivalent BA.4-5 booster was also recommended as part of the spring booster programme in England, very few doses were given, and we were unable to estimate vaccine effectiveness.

The Sanofi/GSK booster was delivered first, with those living in care homes being prioritised for vaccination (15). Those living in care homes may be older and effectiveness may be expected to be lower in these individuals so in addition to adjusting for care home resident status in the main analysis, we also undertook sensitivity analyses where we only included those who were not known to be resident in a care home as of March 2023. We did not find a significant difference in effectiveness by manufacturer in either analysis.

We observed an apparent protective effect of booster vaccination with both manufacturers in the first couple of days post vaccination. The lower odds of vaccination in cases compared to controls in the 0 to 2 days prior to testing is likely related to a combination of deferral of vaccination in individuals who were already aware of their COVID-19 positive status from home lateral flow testing and a healthy vaccinee effect in which those with early symptoms of COVID-19 were more unwell than controls and were less likely to be vaccinated. These effects would only be expected to be transient as indicated by the fact that the odd ratio was close to one for the 3 to 8 day period. The true protective effect of booster vaccination appeared to peak at 9 to 13 days post vaccination. Previously, we have found that the effectiveness of booster vaccination against hospitalisation wanes over time (5). Here we observed also observe evidence of waning, this did not differ significantly by manufacturer although the point estimate beyond 10 weeks after vaccination was lower for the Sanofi/GSK vaccine. It is important to note that the vaccines were used over slightly different time periods, with the Sanofi vaccine being the predominant one used in the first weeks of the programme and so this observation may be confounded by time. Further follow-up would be needed to fully assess waning over time of the Sanofi/GSK and Pfizer BA.4-5 booster vaccines.

The TNCC study design has been widely used to assess influenza vaccine effectiveness, and since the emergence of COVID-19 it has also been used by many groups to assess vaccine effectiveness against COVID-19. In comparison with conventional cohort or case-control study designs, a strength of using the TNCC study design to estimate vaccine effectiveness is that it helps to address unmeasured confounders related to differences in health-seeking behaviours and infectious-disease exposure between vaccinated and unvaccinated people. The TNCC study design requires testing to be independent of vaccination status, which is likely to be the case in a hospital setting. However, there are also several limitations to our study. As an observational study, there may have been unmeasured confounders which we were not able to adjust for. Immunity driven by prior infections may affect vaccine effectiveness and most infections have been undocumented since freely available community testing ended in England. In previous studies (5, 8) and here we found that when we do not adjust for past positivity our results are similar to those of the main analyses (estimates changed by less than 2%), suggesting that these missing data were not likely to have caused a large bias. Including those with past infection is most relevant to public health policy as most of the population have now been infected. A further limitation is that our study relied on hospital coding; errors in the coding may have resulted in the inclusion of some non-respiratory cases and controls in our study (4).

Overall, these results provide reassuring evidence that both the adjuvanted Sanofi/GSK (VidPrevtyn Beta) and Pfizer mRNA BA.4-5 booster vaccines provide a substantial boost in protection against hospitalisation with COVID-19 amongst adults aged 75 years and older for at least 10 weeks after vaccination. Further follow up is needed to better understand the duration of boosted protection with these two vaccines.

## Data Availability

This work is carried out under Regulation 3 of The Health Service (Control of Patient Information; Secretary of State for Health, 2002) using patient identification information without individual patient consent as part of the UKHSA legal requirement for public health surveillance and monitoring of vaccines. As such, authors cannot make the underlying dataset publicly available for ethical and legal reasons. However, all the data used for this analysis is included as aggregated data in the manuscript tables and appendix. Applications for relevant anonymised data should be submitted to the UKHSA Office for Data Release at https://www.gov.uk/government/publications/accessing-ukhsa-protected-data.

## Authors’ Contributions

JLB, NA, FCMK and MR conceptualised the study. FCMK, JS and NA developed the methodology. FCMK and JS curated the data. FCMK accessed and verified the data. FCMK conducted the formal analysis, supported by NA. FCMK wrote the original draft of the manuscript. JLB, NA, GD and MR provided supervision. All co-authors reviewed the manuscript and were responsible for the decision to submit the manuscript.

## Ethics Committee Approval

The study protocol was subject to an internal review by the UK Health Security Agency Research Ethics and Governance Group and was found to be fully compliant with all regulatory requirements. As no regulatory issues were identified, and ethical review is not a requirement for this type of work, it was decided that a full ethical review would not be necessary. UKHSA has legal permission, provided by Regulation 3 of The Health Service (Control of Patient Information) Regulations 2002, to process patient confidential information for national surveillance of communicable diseases and as such, individual patient consent is not required to access records.

## References

1. Andrews N, Stowe J, Kirsebom F, Toffa S, Rickeard T, Gallagher E, et al. Covid-19 Vaccine Effectiveness against the Omicron (B.1.1.529) Variant. New England Journal of Medicine. 2022.

2. Kirsebom FCM, Andrews N, Stowe J, Groves N, Chand M, Ramsay M, et al. Effectiveness of the COVID-19 vaccines against hospitalisation with Omicron sub-lineages BA.4 and BA.5 in England. The Lancet Regional Health – Europe. 2022;23:100537.

3. Kirsebom FCM, Andrews N, Stowe J, Toffa S, Sachdeva R, Gallagher E, et al. COVID-19 vaccine effectiveness against the omicron (BA.2) variant in England. The Lancet Infectious Diseases. 2022;22(7):931–3.

4. Stowe J, Andrews N, Kirsebom F, Ramsay M, Bernal JL. Effectiveness of COVID-19 vaccines against Omicron and Delta hospitalisation, a test negative case-control study. Nature communications. 2022;13(1):5736.

5. Kirsebom FCM, Andrews N, Stowe J, Ramsay M, Lopez Bernal J. Duration of protection of ancestral-strain monovalent vaccines and effectiveness of bivalent BA.1 boosters against COVID-19 hospitalisation in England: a test-negative case-control study. The Lancet Infectious Diseases. 2023;0(0).

6. UKHSA. SARS-CoV-2 variants of concern and variants under investigation in England: Technical briefing 36 2022 [Available from: https://assets.publishing.service.gov.uk/government/uploads/system/uploads/attachment_data/file/1054357/Technical-Briefing-36-11February2022_v2.pdf.

7. UKHSA. SARS-CoV-2 variants of concern and variants under investigation in England: Technical briefing 44 2022 [Available from: https://assets.publishing.service.gov.uk/government/uploads/system/uploads/attachment_data/file/1103191/covid-technical-briefing-44-22-july-2022.pdf.

8. Kirsebom FCM, Harman K, Lunt RJ, Andrews N, Groves N, Aziz NA, et al. Vaccine effectiveness against hospitalisation and comparative odds of hospital admission and severe outcomes with BQ.1, CH.1.1. and XBB.1.5 in England. medRxiv. 2023:2023.07.28.23293333.

9. UK Health Security Agency. National Influenza and COVID-19 surveillance report: Week 33 report (up to week 32 data) 2023 [Available from: https://assets.publishing.service.gov.uk/government/uploads/system/uploads/attachment_data/file/1179183/Weekly_Flu_and_COVID-19_report_w33.pdf.

10. Rasmussen M, Møller FT, Gunalan V, Baig S, Bennedbæk M, Christiansen LE, et al. First cases of SARS-CoV-2 BA.2.86 in Denmark, 2023. Eurosurveillance. 2023;28(36):2300460.

11. UKHSA. SARS-CoV-2 variant surveillance and assessment: technical briefing 53 2023 [Available from: https://www.gov.uk/government/publications/investigation-of-sars-cov-2-variants-technical-briefings/sars-cov-2-variant-surveillance-and-assessment-technical-briefing-53.

12. Department of Health & Social Care. JCVI advice on the UK vaccine response to the Omicron variant 2021 [Available from: https://www.gov.uk/government/publications/uk-vaccine-response-to-the-omicron-variant-jcvi-advice/jcvi-advice-on-the-uk-vaccine-response-to-the-omicron-variant.

13. Joint Committee on Vaccination and Immunisation. JCVI updated statement on the COVID-19 vaccination programme for autumn 2022 2022 [Available from: https://www.gov.uk/government/publications/jcvi-updated-statement-on-the-covid-19-vaccination-programme-for-autumn-2022.

14. UK Health Security Agency. COVID-19: the green book, chapter 14a. Immunisation against infectious diseases: UK Health Security Agency,; 2020.

15. Joint Committee on Vaccination and Immunisation. JCVI statement on spring 2023 COVID-19 vaccinations, 22 February 2023 2023 [Available from: https://www.gov.uk/government/publications/spring-2023-covid-19-vaccination-programme-jcvi-advice-22-february-2023/jcvi-statement-on-spring-2023-covid-19-vaccinations-22-february-2023.

16. Mannino S, Villa M, Apolone G, Weiss NS, Groth N, Aquino I, et al. Effectiveness of adjuvanted influenza vaccination in elderly subjects in northern Italy. Am J Epidemiol. 2012;176(6):527–33.

17. Tregoning JS, Russell RF, Kinnear E. Adjuvanted influenza vaccines. Hum Vaccin Immunother. 2018;14(3):550–64.

18. US Food and Drug Administration. Vaccines and related biological products advisory committee January 26, 2023 meeting presentation -COVID-19 evaluation of next generation vaccines 2023 [Available from: https://www.fda.gov/media/164809/download.

19. Andrews N, Tessier E, Stowe J, Gower C, Kirsebom F, Simmons R, et al. Duration of Protection against Mild and Severe Disease by Covid-19 Vaccines. New England Journal of Medicine. 2022.

20. Tessier E, Edelstein M, Tsang C, Kirsebom F, Gower C, Campbell CNJ, et al. Monitoring the COVID-19 immunisation programme through a national immunisation Management system – England’s experience. International Journal of Medical Informatics. 2023;170:104974.

21. NHS Digital. Cohorting as a Service (CaaS) 2022 [Available from: https://digital.nhs.uk/services/cohorting-as-a-service-caas.

22. NHS Digital. Secondary Uses Service (SUS) 2022 [Available from: https://digital.nhs.uk/services/secondary-uses-service-sus.

23. Tenforde Mw Fau -Weber ZA, Weber Za Fau - Natarajan K, Natarajan K Fau - Klein NP, Klein Np Fau - Kharbanda AB, Kharbanda Ab Fau - Stenehjem E, Stenehjem E Fau - Embi PJ, et al. Early Estimates of Bivalent mRNA Vaccine Effectiveness in Preventing COVID-19-Associated Emergency Department or Urgent Care Encounters and Hospitalizations Among Immunocompetent Adults - VISION Network, Nine States, September-November 2022. MMWR Morbidity and mortality weekly report. 2022(1545-861X (Electronic)).

24. Link-Gelles R Fau - Ciesla AA, Ciesla Aa Fau - Roper LE, Roper Le Fau - Scobie HM, Scobie Hm Fau - Ali AR, Ali Ar Fau - Miller JD, Miller Jd Fau - Wiegand RE, et al. Early Estimates of Bivalent mRNA Booster Dose Vaccine Effectiveness in Preventing Symptomatic SARS-CoV-2 Infection Attributable to Omicron BA.5- and XBB/XBB.1.5-Related Sublineages Among Immunocompetent Adults - Increasing Community Access to Testing Program, United States, December 2022-January 2023. MMWR Morbidity and mortality weekly report. 2023(1545-861X (Electronic)).

25. Launay O, Cachanado M, Luong Nguyen LB, Ninove L, Lachatre M, Ben Ghezala I, et al. Immunogenicity and Safety of Beta-Adjuvanted Recombinant Booster Vaccine. The New England journal of medicine. 2022;387(4):374–6.

26. Dayan GH, Rouphael N, Walsh SR, Chen A, Grunenberg N, Allen M, et al. Efficacy of a bivalent (D614 + B.1.351) SARS-CoV-2 recombinant protein vaccine with AS03 adjuvant in adults: a phase 3, parallel, randomised, modified double-blind, placebo-controlled trial. Lancet Respir Med. 2023.

27. Link-Gelles R, Weber ZA, Reese SE, Payne AB, Gaglani M, Adams K, et al. Estimates of Bivalent mRNA Vaccine Durability in Preventing COVID-19-Associated Hospitalization and Critical Illness Among Adults with and Without Immunocompromising Conditions - VISION Network, September 2022-April 2023. MMWR Morbidity and mortality weekly report. 2023;72(21):579–88.

28. Mateo-Urdiales A, Sacco C, Fotakis EA, Del Manso M, Bella A, Riccardo F, et al. Relative effectiveness of monovalent and bivalent mRNA boosters in preventing severe COVID-19 due to omicron BA.5 infection up to 4 months post-administration in people aged 60 years or older in Italy: a retrospective matched cohort study. The Lancet Infectious diseases. 2023.

