## Supplementary Appendix for "Effectiveness of the adjuvanted Sanofi/GSK (VidPrevtyn Beta) and Pfizer-BioNTech (Comirnaty Original/Omicron BA.4-5) bivalent vaccines against hospitalisation amongst adults aged 75 years and older in England, estimated using a test-negative case control study design"

Supplementary Table 1. SUS Acute respiratory illness ICD-10 code list.

| SUS Acute respiratory illness ICD10 code list | |
| --- | --- |
| J04* | Acute laryngitis and tracheitis |
| J09* | Influenza due to identified avian influenza virus |
| J10* | Influenza with pneumonia, other influenza virus identified |
| J11* | Influenza with pneumonia, virus not identified |
| J12* | Viral pneumonia, not elsewhere classified |
| J13* | Pneumonia due to Streptococcus pneumoniae |
| J14* | Pneumonia due to Haemophilus influenzae |
| J15* | Bacterial pneumonia, not elsewhere classified |
| J16* | Pneumonia due to other infectious organisms, not elsewhere classified |
| J17* | Pneumonia in diseases classified elsewhere |
| J18* | Pneumonia, organism unspecified |
| J20* | Acute bronchitis |
| J21* | Acute bronchiolitis |
| J22* | Unspecified acute lower respiratory infection |
| J80* | ARDS (related to respiratory infection) |
| U071, U072 | COVID-19, virus identified and not identified |
| U04* | Severe acute respiratory syndrome (SARS) |

Supplementary Table 2. Descriptive characteristics of cases and controls included the vaccine effectiveness analysis against hospitalisation amongst those aged 75 years and older in England.

|  |  |  | Overall | | Controls | | Cases | |
| --- | --- | --- | --- | --- | --- | --- | --- | --- |
|  |  |  | n | % | n | % | n | % |
| Vaccination Status | Spring booster | Interval | 14,174 | 100% | 11,169 | 79% | 3,005 | 21% |
|  | None |  | 8,484 | 59.9% | 6,334 | 56.7% | 2,150 | 71.5% |
|  | Sanofi | 0-2 days | 90 | 0.6% | 82 | 0.7% | 8 | 0.3% |
|  |  | 3-8 days | 278 | 2.0% | 215 | 1.9% | 63 | 2.1% |
|  |  | 9-13 days | 253 | 1.8% | 212 | 1.9% | 41 | 1.4% |
|  |  | 2-4 weeks | 964 | 6.8% | 808 | 7.2% | 156 | 5.2% |
|  |  | 5-9 weeks | 1,146 | 8.1% | 1015 | 9.1% | 131 | 4.4% |
|  |  | 10+ weeks | 1,058 | 7.5% | 855 | 7.7% | 203 | 6.8% |
|  | Pfizer BA.4/5 | 0-2 days | 70 | 0.5% | 61 | 0.5% | 9 | 0.3% |
|  |  | 3-8 days | 159 | 1.1% | 132 | 1.2% | 27 | 0.9% |
|  |  | 9-13 days | 152 | 1.1% | 136 | 1.2% | 16 | 0.5% |
|  |  | 2-4 weeks | 535 | 3.8% | 483 | 4.3% | 52 | 1.7% |
|  |  | 5-9 weeks | 653 | 4.6% | 567 | 5.1% | 86 | 2.9% |
|  |  | 10+ weeks | 332 | 2.3% | 269 | 2.4% | 63 | 2.1% |
| Gender | Female |  | 6,930 | 48.9% | 5,578 | 49.9% | 1,352 | 45.0% |
|  | Male |  | 7,088 | 50.0% | 5,443 | 48.7% | 1,645 | 54.7% |
|  | Missing |  | 156 | 1.1% | 148 | 1.3% | 8 | 0.3% |
| Age | 75-79 |  | 3,867 | 27.3% | 3,135 | 28.1% | 732 | 24.4% |
|  | 80-84 |  | 3,852 | 27.2% | 2,986 | 26.7% | 866 | 28.8% |
|  | 85-89 |  | 3,468 | 24.5% | 2,695 | 24.1% | 773 | 25.7% |
|  | 90+ |  | 2,987 | 21.1% | 2,353 | 21.1% | 634 | 21.1% |
| Ethnicity | African |  | 18 | 0.1% | 15 | 0.1% | 3 | 0.1% |
|  | Any other Asian background | | 64 | 0.5% | 55 | 0.5% | 9 | 0.3% |
|  | Any other Black background | | 16 | 0.1% | 14 | 0.1% | 2 | 0.1% |
|  | Any other White background | | 658 | 4.6% | 518 | 4.6% | 140 | 4.7% |
|  | Any other ethnic group | | 78 | 0.6% | 61 | 0.5% | 17 | 0.6% |
|  | Any other mixed background | | 46 | 0.3% | 35 | 0.3% | 11 | 0.4% |
|  | Bangladeshi or British Bangladeshi | | 15 | 0.1% | 13 | 0.1% | 2 | 0.1% |
|  | British, Mixed British | | 12,195 | 86.0% | 9,588 | 85.8% | 2,607 | 86.8% |
|  | Caribbean |  | 51 | 0.4% | 45 | 0.4% | 6 | 0.2% |
|  | Chinese |  | 16 | 0.1% | 7 | 0.1% | 9 | 0.3% |
|  | Indian or British Indian | | 189 | 1.3% | 153 | 1.4% | 36 | 1.2% |
|  | Irish |  | 173 | 1.2% | 142 | 1.3% | 31 | 1.0% |
|  | Pakistani or British Pakistani | | 62 | 0.4% | 52 | 0.5% | 10 | 0.3% |
|  | White and Asian | | 7 | 0.0% | 6 | 0.1% | 1 | 0.0% |
|  | White and Black African | | 3 | 0.0% | 3 | 0.0% | 0 | 0.0% |
|  | White and Black Caribbean | | 8 | 0.1% | 7 | 0.1% | 1 | 0.0% |
|  | Missing |  | 575 | 4.1% | 455 | 4.1% | 120 | 4.0% |
| NHS Region | East of England | | 1,371 | 9.7% | 1,116 | 10.0% | 255 | 8.5% |
|  | London |  | 1,575 | 11.1% | 1,303 | 11.7% | 272 | 9.1% |
|  | Midlands |  | 2,794 | 19.7% | 2,161 | 19.3% | 633 | 21.1% |
|  | North East |  | 2,902 | 20.5% | 2,215 | 19.8% | 687 | 22.9% |
|  | North West |  | 1,607 | 11.3% | 1,302 | 11.7% | 305 | 10.1% |
|  | South East |  | 2,211 | 15.6% | 1,723 | 15.4% | 488 | 16.2% |
|  | South West |  | 1,714 | 12.1% | 1,349 | 12.1% | 365 | 12.1% |
| IMD Quintiles | 1 |  | 2,308 | 16.3% | 1,844 | 16.5% | 464 | 15.4% |
|  | 2 |  | 2,525 | 17.8% | 1,998 | 17.9% | 527 | 17.5% |
|  | 3 |  | 2,886 | 20.4% | 2,272 | 20.3% | 614 | 20.4% |
|  | 4 |  | 3,201 | 22.6% | 2,501 | 22.4% | 700 | 23.3% |
|  | 5 |  | 3,212 | 22.7% | 2,521 | 22.6% | 691 | 23.0% |
|  | Missing |  | 42 | 0.3% | 33 | 0.3% | 9 | 0.3% |
| Previously positive | None |  | 10,712 | 75.6% | 8,142 | 72.9% | 2,570 | 85.5% |
|  | Wild-type |  | 266 | 1.9% | 218 | 2.0% | 48 | 1.6% |
|  | Alpha |  | 312 | 2.2% | 276 | 2.5% | 36 | 1.2% |
|  | Delta |  | 289 | 2.0% | 239 | 2.1% | 50 | 1.7% |
|  | Omicron (before 1st April 2022) | | 996 | 7.0% | 853 | 7.6% | 143 | 4.8% |
|  | Omicron (from 1st April 2022 onwards) | | 1,599 | 11.3% | 1,441 | 12.9% | 158 | 5.3% |
| Risk Status | Carehome resident | | 1,559 | 11.0% | 1,343 | 12.0% | 216 | 7.2% |
|  | CaaS autumn booster cohort | | 13,520 | 95.4% | 10,645 | 95.3% | 2,875 | 95.7% |
|  | CEV |  | 6,519 | 46.0% | 5,174 | 46.3% | 1,345 | 44.8% |
|  | Severely immunosuppressed | | 1,492 | 10.5% | 1,135 | 10.2% | 357 | 11.9% |
| Influenza vaccine 2022/23 | No |  | 1,344 | 9.5% | 1,073 | 9.6% | 271 | 9.0% |
|  | Yes |  | 12,830 | 90.5% | 10,096 | 90.4% | 2,734 | 91.0% |

Supplementary Figure 1. The distribution of cases and controls in the study period over time.

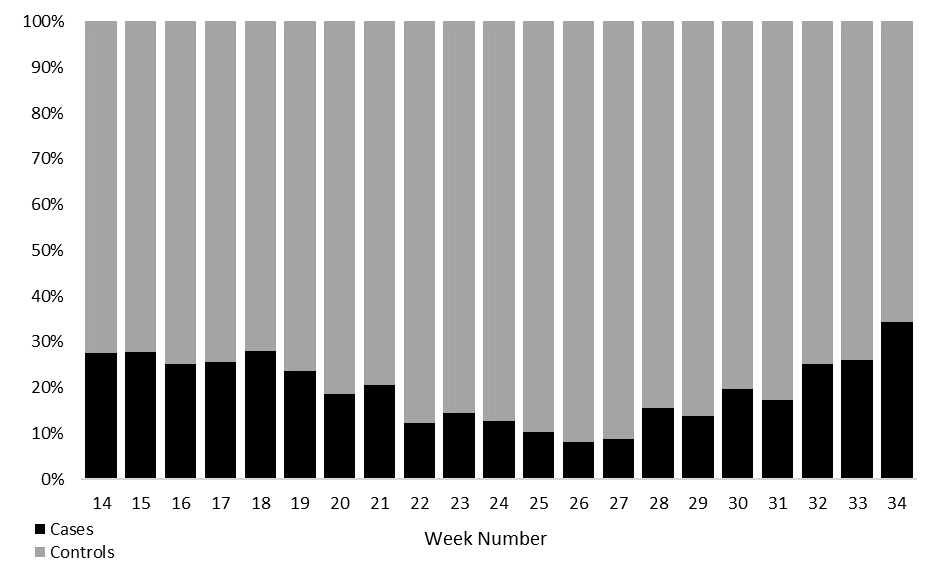

Supplementary Figure 2. The distribution of time since vaccination by manufacturer for cases and controls in the study period.

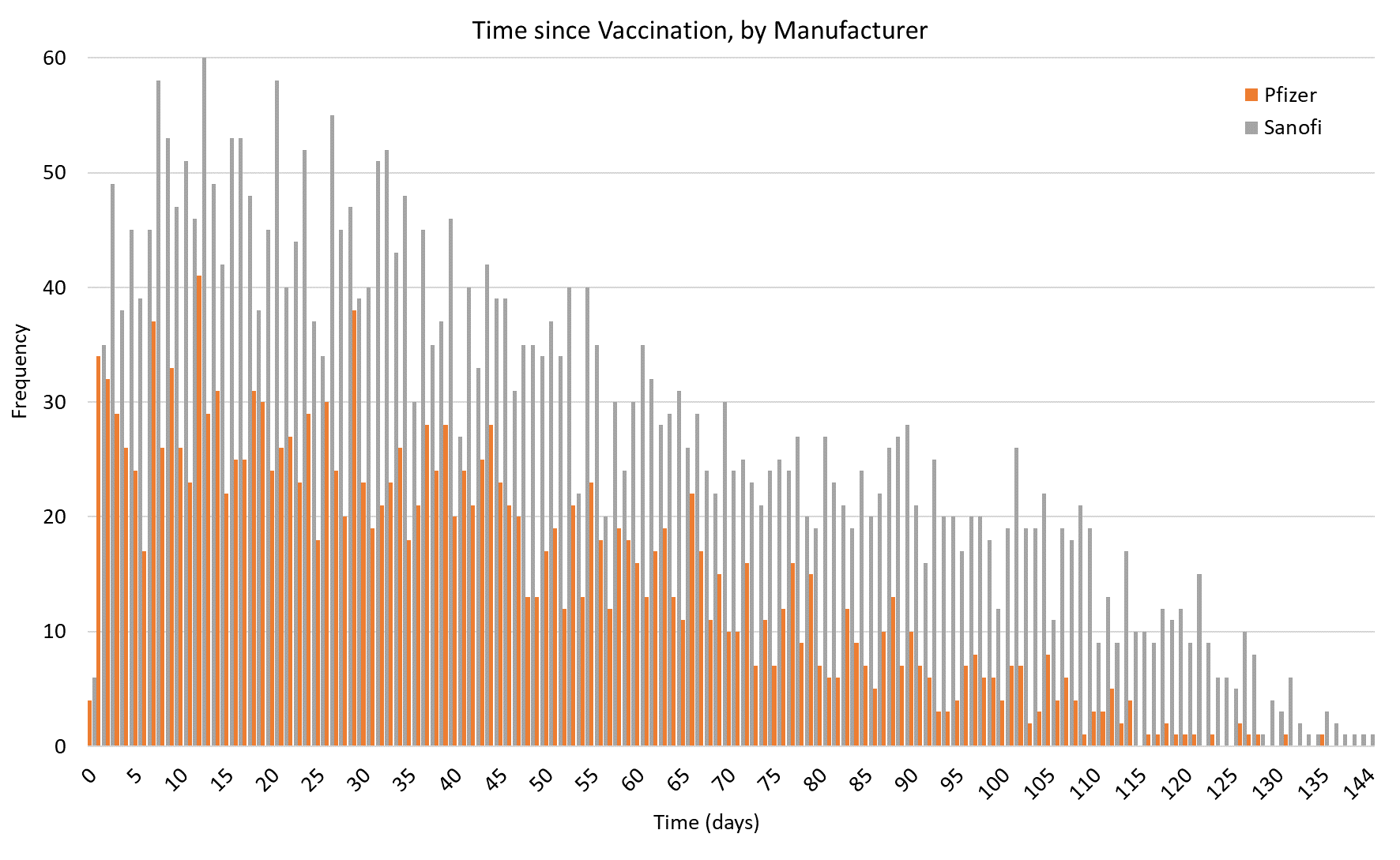
